# Costs of influenza-associated healthcare: an umbrella review and meta-regression analysis

**DOI:** 10.64898/2026.10.08.26365049

**Authors:** Lucy Goodfellow, George Young, Edwin van Leeuwen, Philipp Lambach, Mark Jit, Rosalind M Eggo, Simon R Procter

## Abstract

**Background:** Estimates of the healthcare costs associated with influenza treatment are essential for cost-effectiveness analyses that inform preventative and treatment strategies such as vaccination and non-pharmaceutical interventions. Existing evidence varies across treatment settings, age groups, and countries and there is little data from low- and middle-income settings.

**Methods:** We conducted an umbrella review of systematic reviews and meta-analyses reporting direct admitted and non-admitted hospitalisation costs of seasonal influenza treatment in children, adults, and older people. We supplemented an evidence gap with a systematised review of studies reporting adult non-admitted treatment costs. Costs were extracted and adjusted to 2023 USD. We fitted a Bayesian mixed-effects model to estimate influenza treatment costs using national economic indicators, age group, and treatment setting as *a priori* predictors. We predicted admitted and non-admitted treatment costs for children, adults, and older people in 195 countries and territories.

**Results:** Seven reviews were included in the umbrella review, supplemented by 17 primary studies for adult non-admitted care. Extracted data points most frequently described admitted treatment costs and came from high-income countries. The best-fitting model used healthcare expenditure per capita to predict treatment costs, with country-level random effects. Estimated costs increased with healthcare expenditure per capita, were higher for admitted than non-admitted care, and were highest for adults and lowest for children.

**Conclusions:** This study provides novel estimates of influenza-associated healthcare costs across age groups and treatment settings by synthesising the available global evidence. By extrapolating to settings without data, our estimates can inform future economic evaluation studies of influenza interventions worldwide.

## Introduction

Understanding the economic costs of seasonal influenza is essential for informing cost-effectiveness analyses that guide public health decision-making, including the implementation of vaccine programs and non-pharmaceutical interventions (NPIs) [1,2]. Reliable estimates of the costs associated with influenza-related healthcare are central to such analyses. These costs can vary considerably depending on the treatment setting - admitted or non-admitted hospital treatment, also referred to as inpatient or outpatient treatment - as well as by the age of patients and the country in which care is delivered [3].

Treatment costs have been estimated and used in cost-effectiveness analyses in several countries [4,5], however high-income countries (HICs) are overrepresented in these studies and there is a lack of data from low- and middle-income countries (LMICs), where resource constraints are often greatest [3].

Existing data have been synthesized in various systematic reviews and meta-analyses (SRMAs), often specific to a national income level, age group, or treatment setting [6–9]. Despite these previous analyses, there remain gaps in the evidence; in particular there has been no comprehensive systematic synthesis of data on influenza healthcare costs across settings and age groups.

In this study, we address this gap by conducting an umbrella review of existing SRMAs on the costs of influenza-associated healthcare in children, adults, and older people. Based on identified evidence gaps we supplemented this with a systematised review to address gaps in the literature for estimates of adult non-admitted care costs, for which no previous SRMA was identified. We then fitted a regression model to estimate the costs of influenza-associated admitted and non-admitted care across all age groups in 195 countries and territories (hereon referred to as countries), using national-level health care expenditure per capita as a predictor. Our findings provide globally comparable estimates that can be used to inform future economic evaluations of influenza prevention and treatment programs.

## Methods

### Umbrella review

We performed an umbrella review to compile a list of SRMAs that reported either admitted or non-admitted treatment costs of influenza treatment for children, adults, or older people. We searched Medline and Embase from inception (1946 and 1947, respectively) until 18th October 2024 for systematic reviews using Ovid search engine (full search strategy shown in Table S1). There were no geographical restrictions on our search. We included only SRMAs which contained references to both influenza and cost data in their titles. Two reviewers (LG and GY) independently screened the search results, which were de-duplicated prior to screening. We initially assessed the eligibility of each SRMA based on titles and abstracts to form a shortlist of studies for potential inclusion. We subsequently used this set of potentially eligible studies for full-text screening to generate a final list of included studies.

We extracted primary data from the included SRMAs, restricted to primary data where some degree of age specification was explicit, i.e. not those which had single, unstratified values for the entire population. Where age was reported numerically, we interpreted age ranges in which the midpoint was below 20 as children, between 20 and 55 as adults and above 55 as older people. Data which were simply labelled as ‘>16’ were included in our ‘adults’ category. We did not include data where costs were provided as intervals with no central estimates, or data which were disaggregated into stratifications of influenza cases, such as severity, unless the corresponding proportions of each case type values were reported in which case these proportions were used to generate a weighted cost estimate. We extracted direct medical costs of treatment, and did not include any non-medical costs associated with treatment.

We recorded the countries in which the primary studies were conducted, the currency used, and the currency years in which the cost data were presented. The final data set consisted of cost values corresponding to one of the six combinations of age (children, adults and older people) and treatment setting (admitted and non-admitted).

### Systematised review

Our umbrella review identified only one SRMA with a single primary study that reported costs of non-admitted treatment in an adult population. To supplement this evidence, we conducted a separate systematised review, searching the Medline and Embase databases for studies which contained data specifically on cost of adult non-admitted influenza treatment (full search strategy shown in Table S1). The same age category definitions were used as previously described for the umbrella review. Similarly to the umbrella review, two reviewers (LG and GY) screened the search results after de-duplication. We initially assessed studies’ eligibility based on titles and abstracts to form a shortlist, and subsequently used this set of studies for full-text screening to generate a final list of included studies. We extracted data from this final set of studies to form a dataset containing costs for adult non-admitted influenza treatment. We recorded the countries in which the studies were conducted, the currency used, and the currency years in which the cost data were presented.

### Meta-regression

We adjusted the cost data found in the umbrella review and our systematised review using exchange rates and inflation rates from the World Bank using the *WDI* R package [10,11]. We defined the data year to be the year from which the data was taken, the reported cost year to be the year in which the reported costs were presented (with respect to inflation), and the reported currency to be the currency in which costs were reported (89% of data points were reported in USD). Costs were inflated to 2023 USD using the US GDP deflator. For costs where the reported currency was not USD, and when the data year was earlier than the cost year, we first deflated in local currency and converted to USD using period exchange rates before reinflating to 2023 USD. For any data points where the data year was not given, we assumed that the data year was the same as the reported cost year.

Given that healthcare costs correlate strongly with national-level indicators including gross domestic product per capita (GDPpc) and health care expenditure per capita (HCEpc), we used these two indicators to extrapolate estimates to countries with limited or no data. Specifically, we tested both national GDPpc and HCEpc as potential predictors in the meta-regression analysis (both reported in 2023 USD).

We predicted mean treatment costs based on the natural logarithm of either GDPpc or HCEpc using the *brms* R package [12], which fits Bayesian Regression Models using Markov Chain Monte Carlo (MCMC) methods. We used a Gamma response distribution with a log-link function to ensure non-negative cost estimates. We tested several regression formulae which encompassed varying levels of interaction between the explanatory variables, age group (children, adults, and older people), and treatment settings (admitted and non-admitted) (Table S2). We assumed that there were random effects at the country-level, and also tested formulae which did not include age group. We compared the models using leave-one-out (LOO) cross-validation, and selected the best model based on the largest expected log-predictive density.

We estimated mean treatment costs, with 95% credible intervals (CrIs), using the posterior predictive distribution of the preferred model for each age group and treatment setting in the 195 countries with the necessary predictors provided by the *WDI* R package. For countries with reported cost data, predictions incorporated both global fixed (population-level) effects and country-specific random effects, conditioning on the observed data to produce country-adjusted estimates. For countries without reported cost data, random effects could not be estimated; predictions therefore relied on population-level effects only, marginalising over the random-effects distribution. Here, we assumed that countries with no reported cost data follow the average relationship between the predictor and treatment costs estimated across the observed countries.

## Results

### Reviews

Through the umbrella review search, we identified 116 studies, 47 of which were removed during de-duplication. We reduced the resulting list of 69 studies to 8, following title and abstract screening, and chose 7 studies after full-text screening (Figure 1). During the follow-up supplementary systematised review, we identified 178 studies. We had 105 remaining after de-duplication, and 38 after title and abstract screening. Full-text screening resulted in a final list of 17 primary studies which contained data on cost of adult non-admitted treatment for influenza (Figure 1).

**Figure 1.**
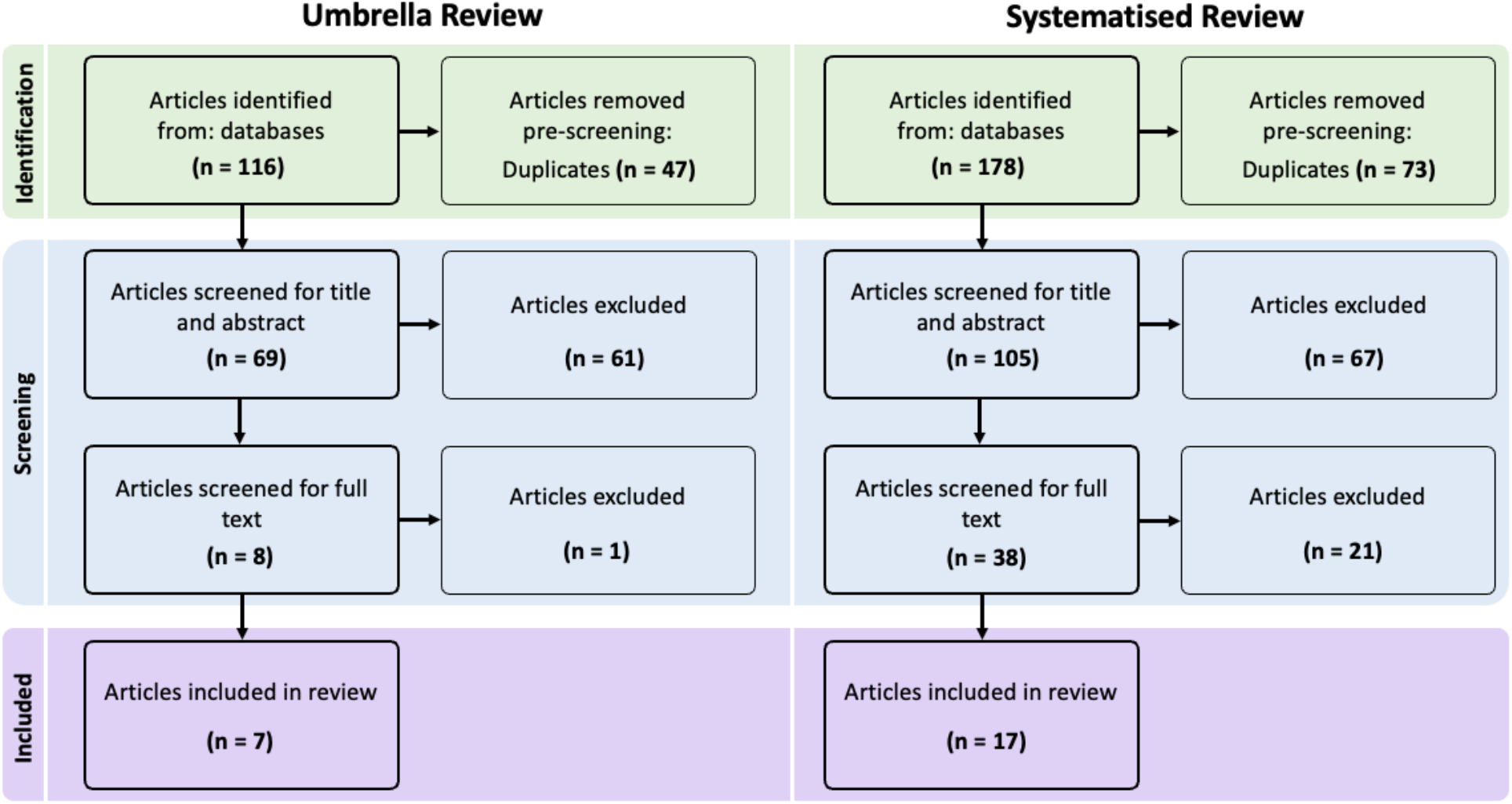
Flowcharts of included/excluded studies in the umbrella and systematised reviews.

The geographical scope of the review studies varied: two of the seven restricted their analysis to LMICs, one to HICs, one to a pre-defined list of countries, one specifically to Thailand, and two did not have restrictions and presented a global analysis (Table 1). 68% of the 140 data entries concerned admitted treatment costs; the remaining 32% relating to non-admitted costs. Most of the data also related to either older people or children, with only 13 for adults, of which 9 were from a single SRMA. Most SRMAs did not specify a date limit and searched their respective databases since inception, but the vast majority of primary studies reported by the SRMAs were published since 2010 (130/140).

**Table 1.**
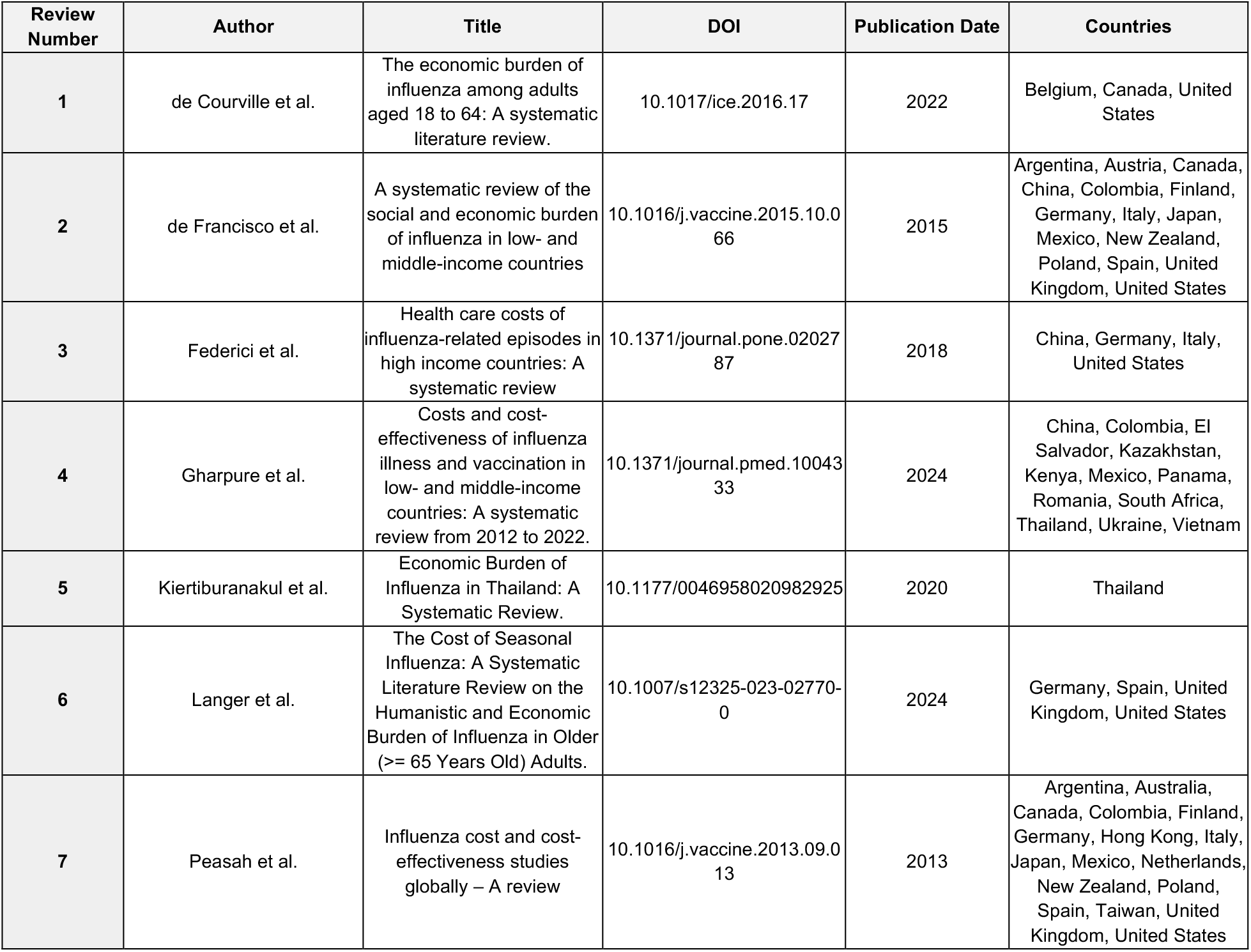
Reviews included in the meta-regression analysis, taken from the umbrella review.

The seven SRMAs each reported substantially varied cost data, with four of the SRMAs including costs that ranged from under $30 to over $10,000. Review 1, which only included costs from HICs (Belgium, Canada, and the United Stated), reported particularly high cost data, ranging from around $7,000 to almost $50,000. This study only contained data on admitted treatment costs for younger adults and older people.

We found only one SRMA with a single primary study specifically for adult non-admitted costs, prompting the systematised review to extract more data for this category. All 17 studies included in the systematised review focused on individual countries. The countries with reported data included in the systematic review were Brazil, China, Germany, Japan, the Netherlands and most commonly the United States, which comprised 11 of the 17 studies (Table 2). A full table of the extracted data from the umbrella and systematic reviews is available online.

**Table 2.** Studies included in the meta-regression analysis, taken from the systematised review. Some studies include multiple cost estimates, for example due to reporting costs over multiple influenza seasons.

| Study number | Author | Title | DOI | Publication year | Country | Currency Year | Currency | Reported costs |
| --- | --- | --- | --- | --- | --- | --- | --- | --- |
| 1 | Nowalk et al. | Cost-effectiveness of recombinant influenza vaccine compared with standard dose influenza vaccine in adults 18-64 years of age. | 10.1016/j.vaccine.2024.07.008 | 2024 | United States | 2020 | USD | 509 |
| 2 | Hu et al. | Economic burden and secondary complications of influenza-related hospitalization among adults in the US: a retrospective cohort study. | 10.1080/13696998.2024.2314429 | 2024 | United States | 2023 | USD | 612-1650 |
| 3 | Vecoso et al. | Cost-Effectiveness Analysis of Influenza A (H1N1) Chemoprophylaxis in Brazil | 10.3389/fphar.2019.00945 | 2019 | Brazil | 2016 | BRL | 12 |
| 4 | Nguyen et al. | The epidemiological and economic impact of a cell-based quadrivalent influenza vaccine in adults in the US: A dynamic modeling approach. | 10.3390/vaccines9101095 | 2021 | United States | 2013 | USD | 158-190 |
| 5 | Nakagawa et al. | A Cost-Effectiveness Analysis of Neuraminidase Inhibitors for Influenza Virus Infections in an Adult-Outpatient Setting in Japan. | 10.1016/j.vhri.2020.07.579 | 2021 | Japan | 2016 | JPY | 13366 |
| 6 | Zeevat et al. | Cost-Effectiveness of Quadrivalent Versus Trivalent Influenza Vaccination in the Dutch National Influenza Prevention Program. | 10.1016/j.jval.2020.11.002 | 2021 | Netherlands | 2018 | EUR | 81 |
| 7 | Cai et al. | Cost-effectiveness of the cell-based quadrivalent versus the standard egg-based quadrivalent influenza vaccine in Germany. | 10.1080/13696998.2021.1908000 | 2021 | Germany | 2019 | EUR | 27 |
| 8 | Putri et al. | Economic burden of seasonal influenza in the United States. | 10.1016/j.vaccine.2018.05.057 | 2018 | United States | 2015 | USD | 74 |
| 9 | Yan et al. | US healthcare costs attributable to type A and type B influenza | 10.1080/21645515.2017.1345400 | 2017 | United States | 2009 | USD | 660-794 |
| 10 | Yang et al. | The economic burden of influenza-associated outpatient visits and hospitalizations in China: A retrospective survey. | 10.1186/s40249-015-0077-6 | 2015 | China | 2013 | CHN | 421 |
| 11 | Prosser et al. | Non-traditional settings for influenza vaccination of adults: Costs and cost effectiveness. | 10.2165/00019053-200826020-00006 | 2008 | United States | 2004 | USD | 287-309 |
| 12 | Molinari et al. | The annual impact of seasonal influenza in the US: Measuring disease burden and costs. | 10.1016/j.vaccine.2007.03.046 | 2007 | United States | 2003 | USD | 280-439 |
| 13 | Meltzer et al. | The economic impact of pandemic influenza in the United States: Priorities for intervention. | 10.3201/eid0505.990507 | 1999 | United States | 1995 | USD | 130 |
| 14 | Ehlken et al. | Cost for physician-diagnosed influenza and influenza-like illnesses on primary care level in Germany – results of a database analysis from May 2010 to April 2012 | 10.1186/s12889-015-1885-0 | 2015 | Germany | 2012 | EUR | 45 |
| 15 | Nichol et al. | Cost-benefit analysis of a strategy to vaccinate healthy working adults against influenza | 10.1001/archinte.161.5.749. | 2001 | United States | 1995 | USD | 130 |
| 16 | Bridges et al. | Effectiveness and Cost-Benefit of Influenza Vaccination of Healthy Working Adults | 10.1001/jama.284.13.1655 | 2000 | United States | 1999 | USD | 106 |
| 17 | Lee et al. | Economic Analysis of Influenza Vaccination and Antiviral Treatment for Healthy Working Adults | 10.7326/0003-4819-137-4-200208200-00005 | 2002 | United States | 2001 | USD | 27 |

### Meta-regression

The best-performing model used the natural logarithm of HCEpc as the predictor of treatment cost, with interaction terms between HCEpc and both treatment setting and age group, and random slopes (LOO comparison results shown in Table S3). Using this model, the absolute residual values increased as the fitted values increased, but without a bias in a positive or negative direction (Figure S3); the relative residuals remained fairly constant as fitted values increased.

Estimated mean treatment costs increased with healthcare expenditure per capita in both treatment settings and all age groups (Figure 2). The data points from the literature were typically, but not consistently, captured within the 95% credible interval of the predictive model; by incorporating country-level fixed effects where data is available, we account for these “outlier” costs in our national-level estimates where possible (all estimated costs are shown in Table S4). Estimated treatment costs were consistently highest for adults, and lowest for children, but that these costs did not significantly differ for fixed HCEpc (Figure S1).

**Figure 2.**
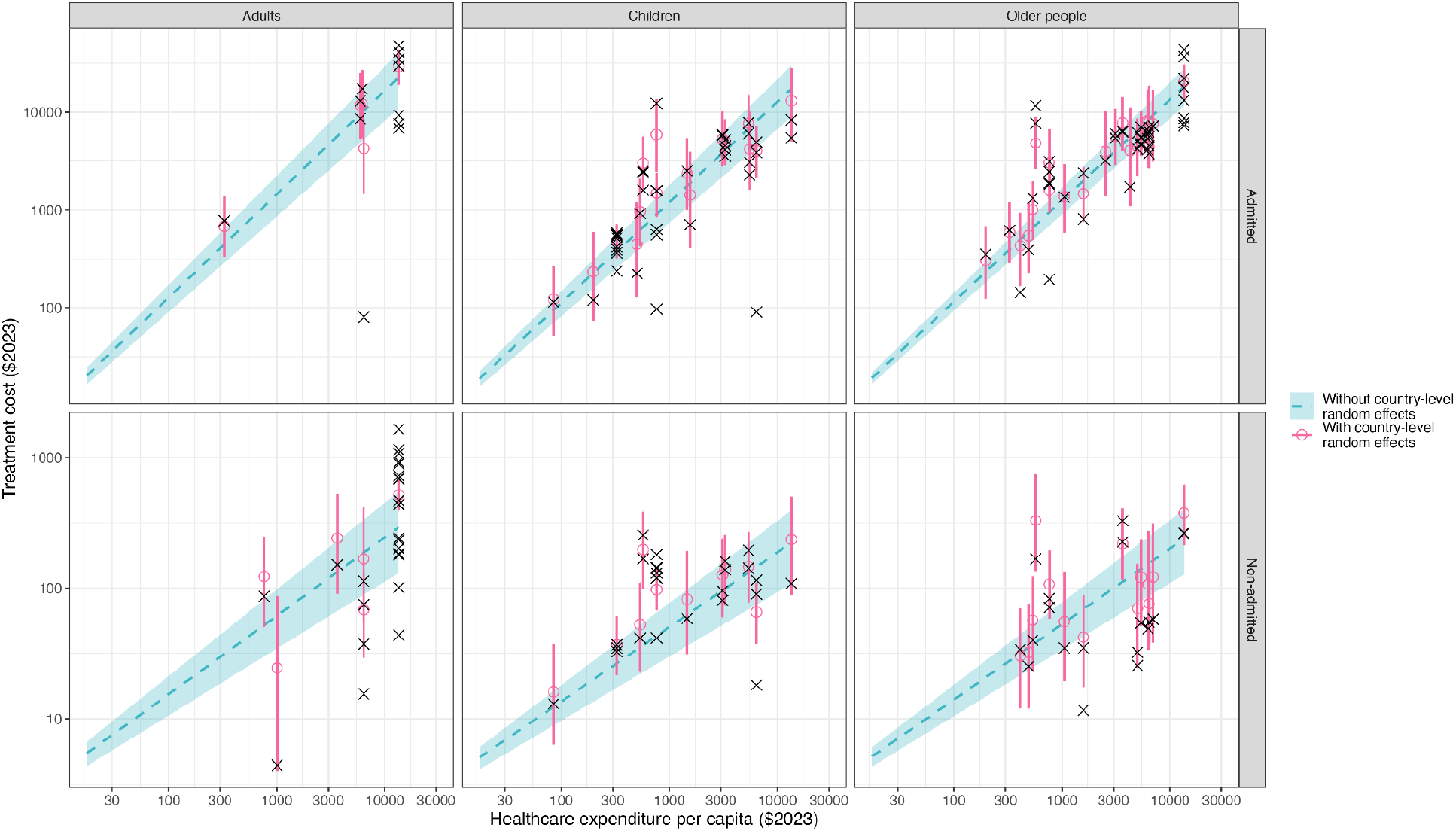
Observed (crosses) and predicted costs of treatment (with 95% credible intervals), by healthcare expenditure per capita, on a log-log scale, for each age group and treatment setting. Dotted line represents the mean cost estimate with no country-level random effects included, with the blue ribbon showing the 95% credible interval of the mean. Mean cost estimates with country-level random effects included are shown in pink, with 95% credible intervals of the mean.

We found that the predicted costs of treatment in each setting and broad age group differed widely across WHO regions, where the lowest costs were found in the African Region and the highest in the European Region and the Region of the Americas (Figure 3). These differences are largely driven by regional differences in HCEpc. Within each region, there was still large variation in the expected mean cost of influenza treatment, indicating that using a regional estimate (or data from a nearby country) is likely insufficient to estimate treatment costs due to within-region differences and nuances better captured by HCEpc.

**Figure 3.**
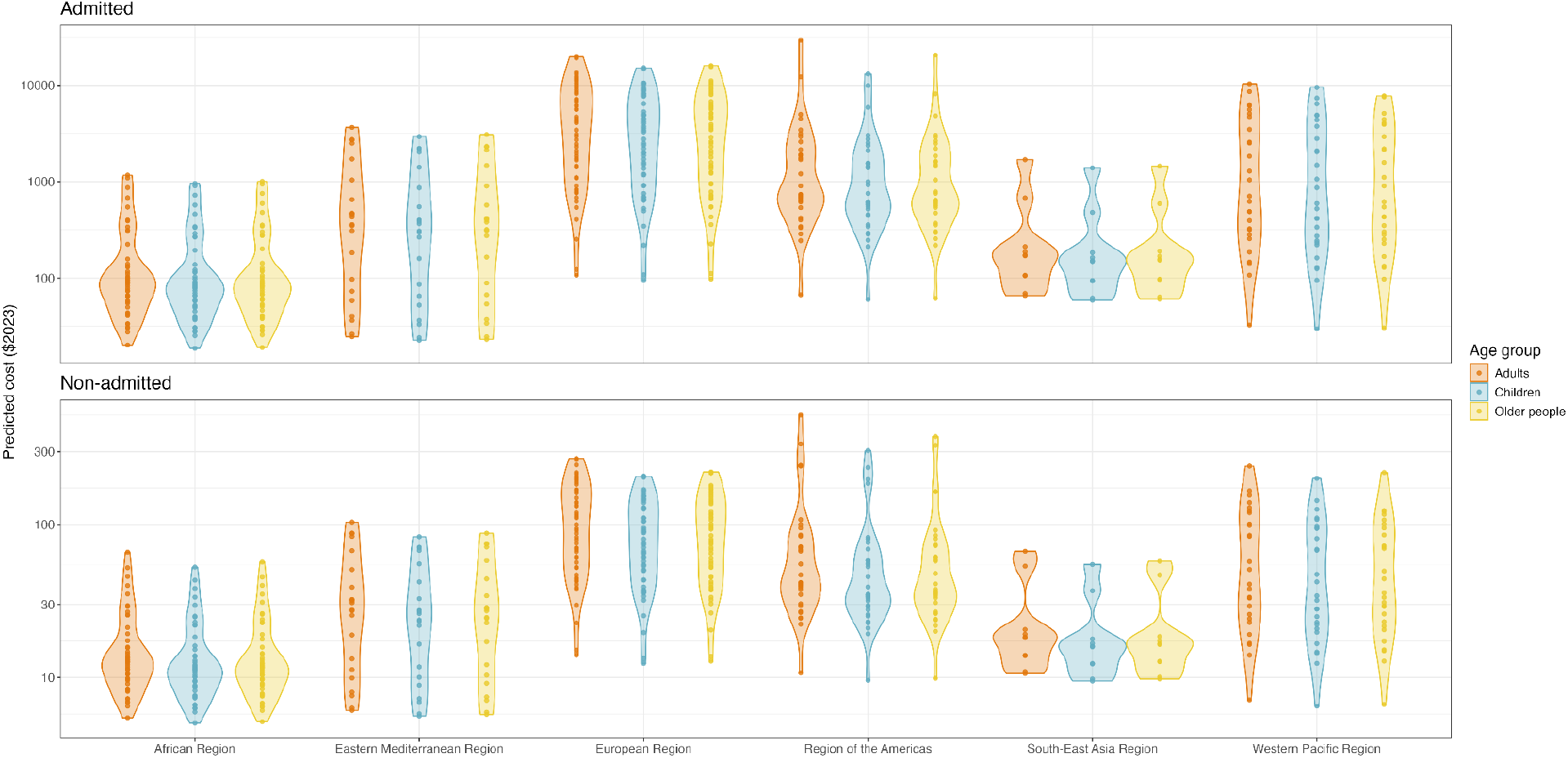
Predicted mean influenza treatment costs in each WHO region, on a log scale, for each age group and treatment setting.

## Discussion

To our knowledge, this is the first study synthesising and predicting global influenza treatment costs by using a meta-regression model which effectively combines countries with large amounts of reported data with those with very little reported data. We conducted two reviews to collate evidence on the costs of influenza treatment across the globe, and categorised data into values for admitted and non-admitted influenza treatment.

We found that healthcare expenditure per capita was the best-fitting predictor of influenza treatment costs, which increased with healthcare expenditure per capita. Admitted care was typically associated with higher treatment costs than non-admitted care. Adults were associated with the highest treatment costs in both settings, while children were associated with the lowest treatment costs, but differences between age groups were not substantially different considering the uncertainty around estimates. We found that estimated mean influenza treatment costs varied substantially between and within global regions; while mean admitted costs for older people ranged from $19 - $1,007 in the African Region, and $5 - $57 for non-admitted treatment, these treatment costs ranged from $62 - $20,633 and $10 - $377, respectively, in the Region of the Americas.

Our meta-regression allowed us to formally predict the cost of influenza treatment for 195 countries around the world, allowing for international and cross-region comparisons of the relative costs of treatment in admitted and non-admitted settings, and serving as a database for future costing and modelling studies on influenza. Many such studies currently rely on estimates which are based on sparse data or unfounded assumptions and are therefore lacking accuracy. Offering values following the aggregation of data taken from many different studies in a range of settings therefore has the potential to improve the accuracy of future work, especially in LMICs, where there is a general lack of existing healthcare cost data. Dividing the data into distinct age and treatment setting categories and producing separate predictions for each category increases the versatility of the results in their application to future studies. By considering various predictive models, we evaluated the best-fitting relationships between age group, treatment setting, and healthcare expenditure per capita.

In this analysis, we included only direct costs of influenza, omitting indirect costs such as travel and loss of income due to time off work, as the data for these aspects are scarce. Costs are differentially borne by governments and by individuals in different countries. Further, the estimates of mean treatment costs produced here largely do not account for national-level nuances in influenza treatment, with the exception of countries with observed data where we could apply both fixed and random effects. The data used in our study is somewhat limited by its skew towards HICs, particularly the USA, which is reflective of a wider bias towards richer settings present in the literature amongst influenza studies.

Comparability between data sources may also be affected by differences in seasonal influenza severity between study periods; this could not be accounted for in the analysis. Despite this, the model outputs presented here clearly show that there is still adequate data from LMICs, and the presence of data from all six inhabited continents demonstrates the global representativeness of this study.

## Conclusion

We produced cost predictions for treating children, adults, and older people in both admitted and non-admitted care, increasing the utility of our results for future research. While the majority of the data were taken from HICs, they spanned a large range of income groups and were taken from a vast range of geographical regions. The work presented here can be used in future by economic evaluation studies of influenza interventions around the globe to improve accuracy where data has not been previously available, which will be especially useful in LMICs where resources are often highly constrained.

## Acknowledgements

LG, MJ, RME, and SRP were funded through the Task Force for Global Health (grant numbers INF-CDC-R2R; INF-CDC-PV3, INF-CDC-PV4) in collaboration with Partnership for Influenza Vaccine Introduction, Ready2Respond, Wellcome Trust, and Centers for Disease Control and Prevention. LG, MJ, RME, and SRP were funded by the World Health Organization (grant number 2305-IAI-PDR-Flu-Vac). RME was supported by the Medical Research Council (MR/X033260/1), by the NIHR Health and Social Care Delivery Research programme (NIHR158218) and by the National Institute for Health and Care Research (NIHR) Health Protection Research Unit in Health Analytics & Modelling, a partnership between the UK Health Security Agency, Imperial College London and LSHTM (grant code NIHR207404). The views expressed are those of the author(s) and not necessarily those of the NIHR, UK Health Security Agency or the Department of Health and Social Care. P Lambach works for the World Health Organization (WHO). The authors alone are responsible for the views expressed in this publication and they do not necessarily represent the decisions, policy or views of the WHO. The funders had no other role in study design, data collection and analysis, decision to publish, or preparation of the manuscript.

## Data availability

All code and data used in this study are available from https://github.com/lucy-gf/treatment_cost_analysis.

