## Supplementary Materials for "Costs of influenza-associated healthcare: an umbrella review and meta-regression analysis"

| Umbrella Review | #1 | (Flu OR influenza).m_titl. |
| --- | --- | --- |
|  | #2 | (Cost OR economic$).m_titl. |
|  | #3 | #1 AND #2 |
| Systematised Review | #1 | (Flu OR influenza).m_titl. |
|  | #2 | (Cost OR economic$).m_titl. |
|  | #3 | Outpatient$.ab. |
|  | #4 | #1 AND #2 AND #3 |

**Table S1.** Search strategies used in Medline via OVID.

| **Model description** | **Model formula** |
| --- | --- |
| No interaction, random intercept | cost ~ 0 + log(*var*) + treatment_type + age_group + (1 \| ISO3C) |
| No interaction, random slope | cost ~ 0 + log(*var*) + (1 + treatment_type + age_group \| ISO3C) |
| Interaction, random intercept | cost ~ 0 + log(*var*):treatment_type + log(*var*):age_group + (1 \| ISO3C) |
| Interaction, random slope | cost ~ 0 + log(*var*):treatment_type + log(*var*):age_group + (1 + treatment_type + age_group \| ISO3C) |
| Interaction, random slope, without age | cost ~ 0 + log(*var*):treatment_type + (1 + treatment_type \| ISO3C) |

**Table S2.** Formulae tested, where *var* is either healthcare expenditure per capita or gross domestic product per capita.

| **Model** | **ELPD Difference** |
| --- | --- |
| HCE_interaction_rand_slope | 0 |
| HCE_interaction_rand_slope_no_age | -1.22 |
| GDP_interaction_rand_slope | -1.74 |
| GDP_interaction_rand_slope_no_age | -3.38 |
| HCE_interaction_rand_intercept | -4.2 |
| GDP_interaction_rand_intercept | -5.3 |
| GDP_no_interaction_rand_slope | -11.31 |
| HCE_no_interaction_rand_intercept | -13.6 |
| GDP_no_interaction_rand_intercept | -13.74 |
| HCE_no_interaction_rand_slope | -13.81 |

**Table S3.** Differences in expected log-predictive density (ELPD) across each model used in the meta-regression, where the best-performing model has ELPD difference 0 and all worse-performing models have a negative ELPD difference. The best-performing model used the formula: cost ~ 0 + log(HCEpc):treatment_type + log(HCEpc):age_group + (1 + treatment_type + age_group | ISO3C).

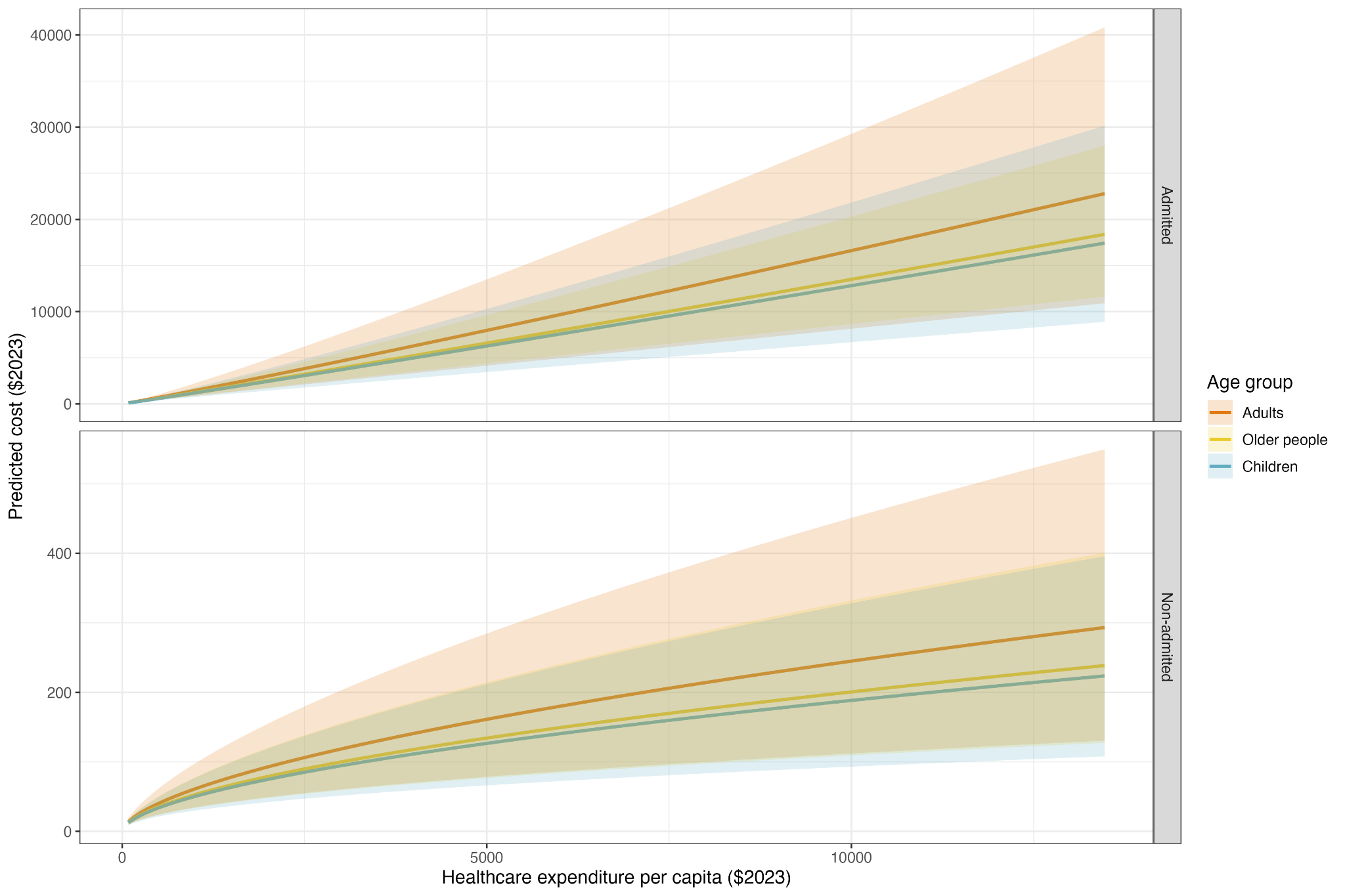

**Fig S1.** Predicted mean treatment costs, and 95% credible intervals of the mean, by healthcare expenditure per capita, for each broad age group and treatment setting.

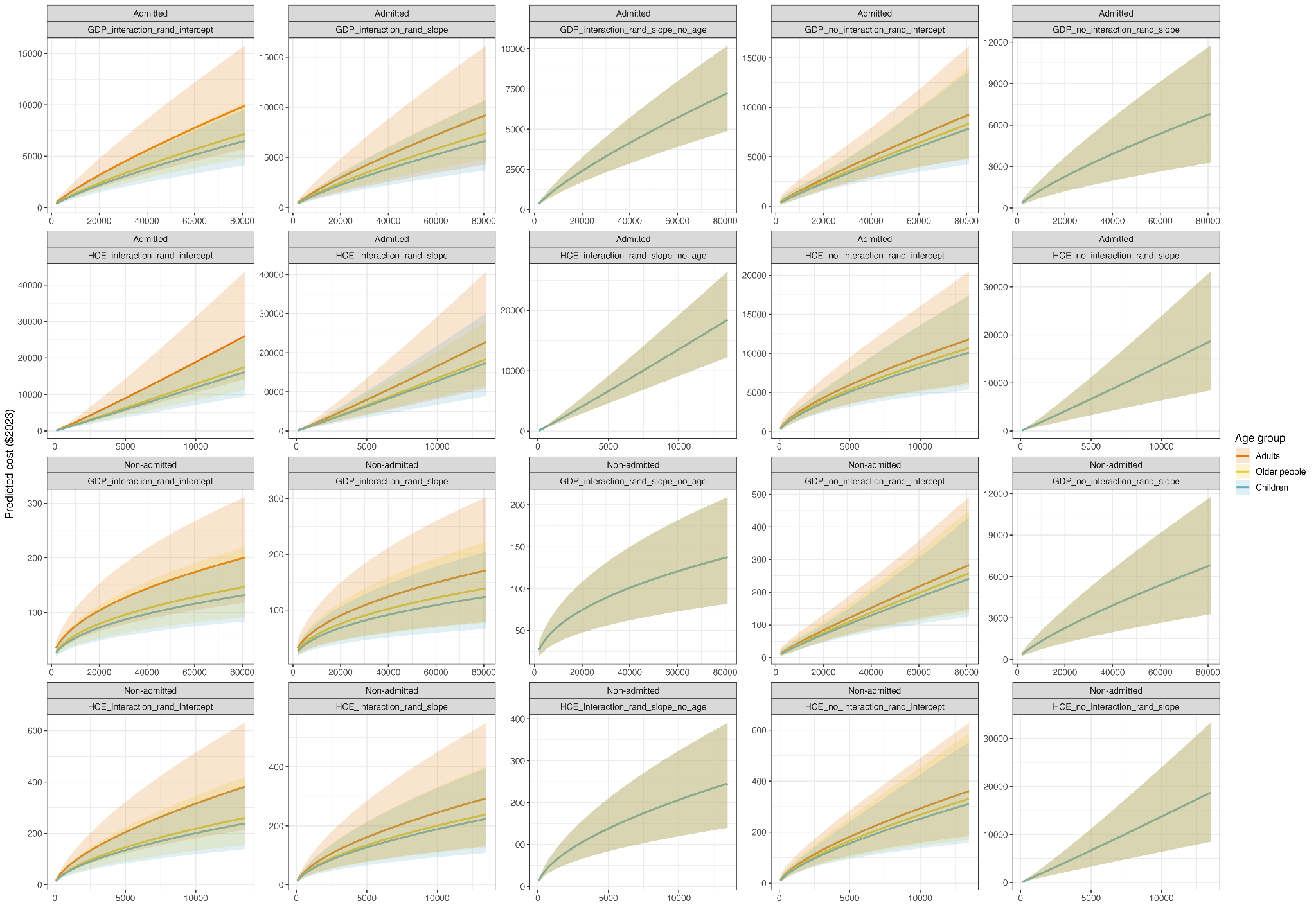

**Fig S2.** Predicted treatment costs, and 95% credible intervals, by healthcare expenditure per capita or gross domestic product per capita, as predicted by each specified model, for each broad age group and treatment setting.

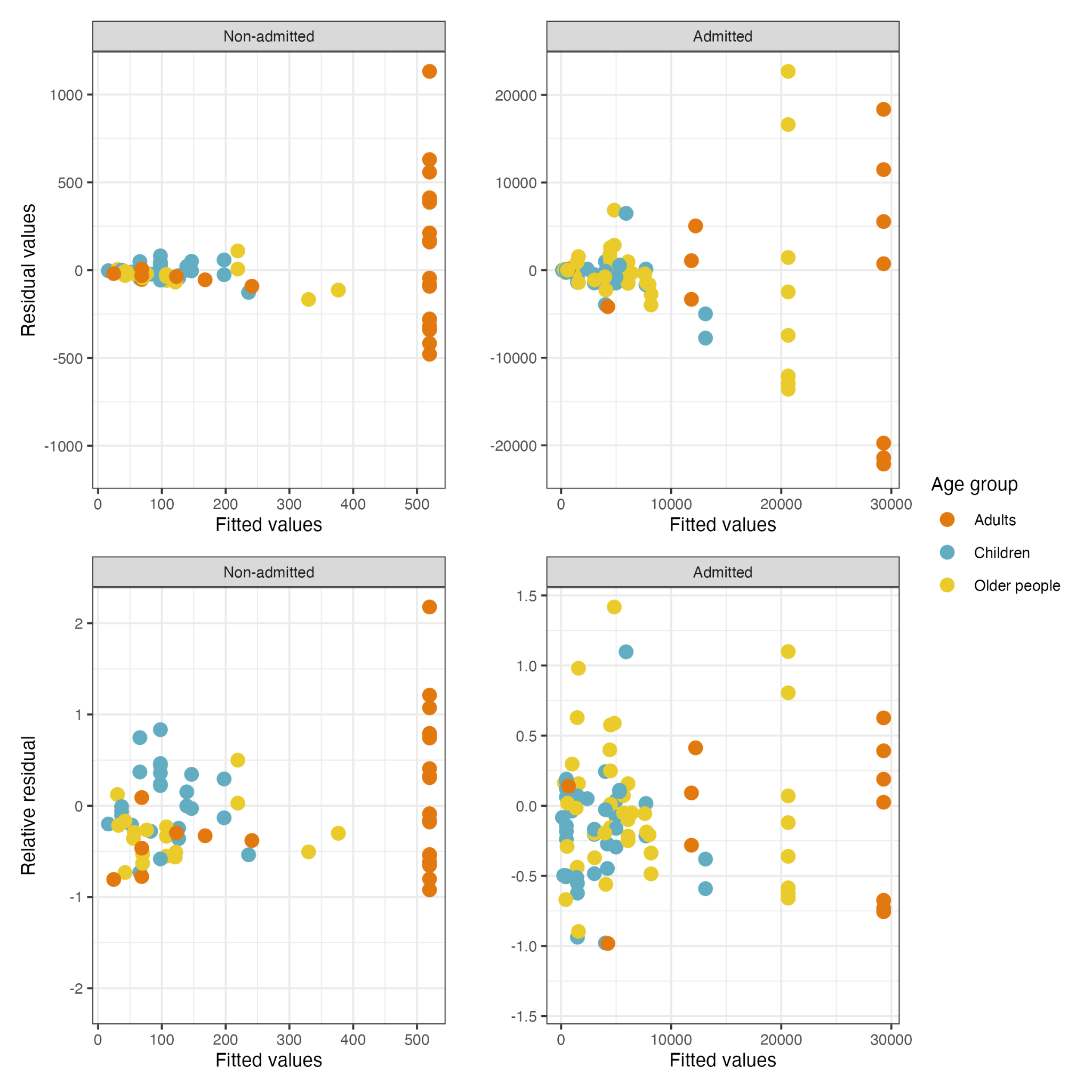

**Fig S3.** Bayesian regression model fitted values vs residuals for the best predictive model, and relative residuals (residual value divided by fitted value).

| **ISO Code** | **Country** | **Healthcare expenditure per capita** | **Admitted treatment (Adults)** | **Admitted treatment (Children)** | **Admitted treatment (Older people)** | **Non-admitted treatment (Adults)** | **Non-admitted treatment (Children)** | **Non-admitted treatment (Older people)** |
| --- | --- | --- | --- | --- | --- | --- | --- | --- |
| MDG | Madagascar | 17.44 | 20.19 (16.37, 24.34) | 18.66 (15.39, 22.22) | 19.05 (16.68, 21.73) | 5.44 (4.33, 6.67) | 5.03 (4.08, 6.04) | 5.14 (4.29, 6.06) |
| SDN | Sudan | 21.08 | 24.65 (19.70, 30.08) | 22.66 (18.45, 27.30) | 23.16 (20.10, 26.66) | 6.09 (4.77, 7.56) | 5.60 (4.48, 6.80) | 5.73 (4.73, 6.83) |
| SOM | Somalia, Fed. Rep. | 22.37 | 26.24 (20.88, 32.14) | 24.09 (19.53, 29.11) | 24.62 (21.31, 28.41) | 6.31 (4.92, 7.86) | 5.79 (4.62, 7.06) | 5.93 (4.88, 7.09) |
| COD | Congo, Dem. Rep. | 23.43 | 27.55 (21.84, 33.85) | 25.26 (20.41, 30.61) | 25.83 (22.30, 29.87) | 6.49 (5.03, 8.11) | 5.95 (4.72, 7.27) | 6.09 (4.99, 7.30) |
| GMB | Gambia, The | 25.74 | 30.43 (23.95, 37.60) | 27.82 (22.34, 33.90) | 28.46 (24.47, 33.05) | 6.86 (5.28, 8.63) | 6.27 (4.95, 7.72) | 6.43 (5.24, 7.75) |
| NER | Niger | 26.01 | 30.76 (24.19, 38.03) | 28.11 (22.56, 34.29) | 28.77 (24.72, 33.43) | 6.91 (5.31, 8.69) | 6.31 (4.97, 7.77) | 6.47 (5.26, 7.80) |
| LAO | Lao PDR | 27.42 | 32.51 (25.47, 40.34) | 29.67 (23.72, 36.30) | 30.37 (26.03, 35.37) | 7.13 (5.46, 9.00) | 6.50 (5.10, 8.03) | 6.66 (5.41, 8.06) |
| BDI | Burundi | 28.14 | 33.42 (26.13, 41.52) | 30.47 (24.32, 37.34) | 31.20 (26.71, 36.38) | 7.24 (5.53, 9.16) | 6.60 (5.17, 8.16) | 6.76 (5.48, 8.20) |
| ERI | Eritrea | 28.3 | 33.61 (26.27, 41.78) | 30.65 (24.45, 37.57) | 31.38 (26.85, 36.60) | 7.26 (5.54, 9.19) | 6.62 (5.18, 8.19) | 6.79 (5.50, 8.22) |
| PAK | Pakistan | 30.49 | 36.36 (28.25, 45.41) | 33.08 (26.25, 40.73) | 33.89 (28.90, 39.65) | 7.59 (5.76, 9.66) | 6.90 (5.38, 8.58) | 7.08 (5.71, 8.62) |
| SYR | Syrian Arab Republic | 33.34 | 39.95 (30.84, 50.18) | 36.26 (28.60, 44.88) | 37.17 (31.56, 43.67) | 8.01 (6.03, 10.25) | 7.26 (5.62, 9.08) | 7.46 (5.97, 9.12) |
| MLI | Mali | 33.98 | 40.76 (31.42, 51.26) | 36.98 (29.13, 45.81) | 37.90 (32.16, 44.57) | 8.10 (6.09, 10.38) | 7.34 (5.67, 9.19) | 7.54 (6.03, 9.23) |
| ETH | Ethiopia | 34.8 | 41.80 (32.16, 52.64) | 37.89 (29.80, 47.01) | 38.84 (32.92, 45.73) | 8.21 (6.16, 10.54) | 7.44 (5.74, 9.33) | 7.64 (6.11, 9.37) |
| SLE | Sierra Leone | 35.76 | 43.02 (33.03, 54.27) | 38.97 (30.58, 48.43) | 39.96 (33.81, 47.09) | 8.35 (6.25, 10.73) | 7.56 (5.82, 9.49) | 7.76 (6.19, 9.53) |
| TZA | Tanzania | 36.26 | 43.64 (33.47, 55.10) | 39.52 (30.99, 49.15) | 40.53 (34.27, 47.79) | 8.42 (6.30, 10.83) | 7.62 (5.86, 9.57) | 7.82 (6.24, 9.62) |
| MWI | Malawi | 41.27 | 50.02 (37.99, 63.67) | 45.13 (35.07, 56.56) | 46.32 (38.93, 54.94) | 9.09 (6.73, 11.80) | 8.20 (6.24, 10.38) | 8.43 (6.66, 10.43) |
| UGA | Uganda | 45.21 | 55.08 (41.54, 70.51) | 49.57 (38.27, 62.45) | 50.90 (42.59, 60.62) | 9.60 (7.05, 12.54) | 8.64 (6.53, 11.00) | 8.88 (6.98, 11.05) |
| BEN | Benin | 46.53 | 56.77 (42.72, 72.80) | 51.05 (39.34, 64.43) | 52.42 (43.81, 62.52) | 9.77 (7.15, 12.78) | 8.78 (6.62, 11.20) | 9.03 (7.08, 11.25) |
| TCD | Chad | 47.44 | 57.94 (43.53, 74.40) | 52.07 (40.07, 65.80) | 53.48 (44.66, 63.84) | 9.88 (7.22, 12.95) | 8.87 (6.69, 11.33) | 9.13 (7.15, 11.39) |
| YEM | Yemen, Rep. | 47.78 | 58.38 (43.84, 75.00) | 52.46 (40.35, 66.31) | 53.88 (44.97, 64.34) | 9.92 (7.25, 13.01) | 8.91 (6.71, 11.38) | 9.17 (7.18, 11.44) |
| RWA | Rwanda | 52.66 | 64.68 (48.21, 83.59) | 57.96 (44.28, 73.68) | 59.57 (49.49, 71.43) | 10.51 (7.62, 13.88) | 9.42 (7.04, 12.10) | 9.70 (7.54, 12.17) |
| MOZ | Mozambique | 52.96 | 65.07 (48.48, 84.13) | 58.30 (44.52, 74.14) | 59.92 (49.77, 71.88) | 10.55 (7.64, 13.93) | 9.45 (7.06, 12.15) | 9.73 (7.56, 12.21) |
| BGD | Bangladesh | 53.46 | 65.73 (48.93, 85.02) | 58.87 (44.93, 74.91) | 60.51 (50.24, 72.61) | 10.61 (7.68, 14.02) | 9.50 (7.09, 12.22) | 9.78 (7.60, 12.28) |
| CAF | Central African Republic | 53.81 | 66.18 (49.24, 85.64) | 59.26 (45.21, 75.44) | 60.92 (50.56, 73.12) | 10.65 (7.71, 14.08) | 9.53 (7.11, 12.27) | 9.82 (7.63, 12.33) |
| HTI | Haiti | 54.41 | 66.96 (49.78, 86.71) | 59.95 (45.69, 76.35) | 61.62 (51.11, 74.00) | 10.72 (7.75, 14.18) | 9.59 (7.15, 12.35) | 9.88 (7.67, 12.42) |
| MMR | Myanmar | 56.19 | 69.26 (51.37, 89.87) | 61.95 (47.11, 79.06) | 63.70 (52.75, 76.61) | 10.93 (7.88, 14.49) | 9.77 (7.27, 12.61) | 10.07 (7.80, 12.67) |
| AFG | Afghanistan | 59.07 | 73.01 (53.94, 95.03) | 65.21 (49.42, 83.46) | 67.07 (55.41, 80.84) | 11.26 (8.08, 14.98) | 10.05 (7.45, 13.01) | 10.36 (8.00, 13.08) |
| GIN | Guinea | 59.33 | 73.35 (54.18, 95.50) | 65.51 (49.63, 83.86) | 67.37 (55.66, 81.23) | 11.29 (8.10, 15.02) | 10.08 (7.46, 13.05) | 10.39 (8.02, 13.12) |
| TGO | Togo | 59.61 | 73.71 (54.42, 96.00) | 65.82 (49.85, 84.28) | 67.70 (55.91, 81.63) | 11.32 (8.12, 15.07) | 10.10 (7.48, 13.08) | 10.41 (8.03, 13.15) |
| ZWE | Zimbabwe | 62.93 | 78.05 (57.39, 102.00) | 69.59 (52.50, 89.40) | 71.60 (58.98, 86.55) | 11.69 (8.35, 15.62) | 10.42 (7.68, 13.54) | 10.74 (8.26, 13.61) |
| NGA | Nigeria | 66.82 | 83.15 (60.86, 109.10) | 74.01 (55.61, 95.41) | 76.17 (62.57, 92.32) | 12.12 (8.61, 16.25) | 10.78 (7.91, 14.06) | 11.12 (8.52, 14.14) |
| GHA | Ghana | 70.26 | 87.67 (63.91, 115.30) | 77.92 (58.34, 100.70) | 80.22 (65.73, 97.45) | 12.49 (8.84, 16.80) | 11.09 (8.11, 14.51) | 11.45 (8.74, 14.59) |
| BFA | Burkina Faso | 71.73 | 89.61 (65.23, 118.10) | 79.61 (59.51, 103.00) | 81.96 (67.09, 99.66) | 12.64 (8.93, 17.04) | 11.22 (8.19, 14.70) | 11.58 (8.83, 14.78) |
| SSD | South Sudan | 72.76 | 90.96 (66.14, 119.90) | 80.77 (60.32, 104.60) | 83.17 (68.03, 101.20) | 12.75 (8.99, 17.20) | 11.32 (8.25, 14.83) | 11.68 (8.89, 14.92) |
| SEN | Senegal | 73.23 | 91.58 (66.55, 120.80) | 81.31 (60.69, 105.40) | 83.72 (68.47, 101.90) | 12.80 (9.02, 17.27) | 11.36 (8.28, 14.89) | 11.72 (8.92, 14.98) |
| COG | Congo, Rep. | 75.55 | 94.65 (68.62, 125.10) | 83.96 (62.54, 109.00) | 86.47 (70.61, 105.40) | 13.04 (9.17, 17.63) | 11.56 (8.41, 15.19) | 11.93 (9.07, 15.27) |
| AGO | Angola | 76.07 | 95.33 (69.08, 126.10) | 84.55 (62.95, 109.80) | 87.08 (71.08, 106.20) | 13.09 (9.20, 17.71) | 11.61 (8.44, 15.25) | 11.98 (9.10, 15.34) |
| DJI | Djibouti | 77.57 | 97.32 (70.41, 128.80) | 86.26 (64.13, 112.20) | 88.86 (72.46, 108.40) | 13.25 (9.30, 17.94) | 11.74 (8.52, 15.44) | 12.12 (9.19, 15.53) |
| CMR | Cameroon | 79.1 | 99.34 (71.77, 131.70) | 88.01 (65.34, 114.60) | 90.66 (73.87, 110.70) | 13.40 (9.39, 18.18) | 11.87 (8.60, 15.63) | 12.25 (9.28, 15.72) |
| ZMB | Zambia | 79.56 | 99.95 (72.17, 132.50) | 88.53 (65.70, 115.30) | 91.20 (74.29, 111.40) | 13.45 (9.42, 18.25) | 11.90 (8.62, 15.69) | 12.29 (9.31, 15.78) |
| GNB | Guinea-Bissau | 82.14 | 103.40 (74.46, 137.30) | 91.48 (67.74, 119.30) | 94.26 (76.66, 115.30) | 13.71 (9.57, 18.64) | 12.12 (8.76, 16.01) | 12.52 (9.46, 16.10) |
| NPL | Nepal | 83.96 | 105.80 (76.08, 140.70) | 93.56 (69.17, 122.20) | 96.42 (78.33, 118.10) | 13.89 (9.68, 18.91) | 12.28 (8.85, 16.23) | 12.68 (9.57, 16.32) |
| IND | India | 84.69 | 106.80 (76.72, 142.10) | 94.40 (69.75, 123.40) | 97.28 (79.00, 119.20) | 13.96 (9.72, 19.02) | 12.34 (8.89, 16.32) | 12.74 (9.61, 16.41) |
| KEN | Kenya | 84.96 | 139.70 (36.58, 370.10) | 121.80 (51.58, 267.20) | 127.20 (35.94, 334.80) | 19.32 (3.94, 58.97) | 16.10 (6.32, 37.09) | 17.84 (3.69, 55.33) |
| TJK | Tajikistan | 85.15 | 107.40 (77.13, 143.00) | 94.92 (70.11, 124.10) | 97.83 (79.42, 119.90) | 14.01 (9.75, 19.09) | 12.37 (8.92, 16.37) | 12.78 (9.64, 16.47) |
| PNG | Papua New Guinea | 85.15 | 107.40 (77.13, 143.00) | 94.93 (70.11, 124.10) | 97.83 (79.43, 119.90) | 14.01 (9.75, 19.09) | 12.37 (8.92, 16.37) | 12.78 (9.64, 16.47) |
| CIV | Cote d'Ivoire | 87.68 | 110.70 (79.37, 147.70) | 97.82 (72.10, 128.10) | 100.80 (81.75, 123.70) | 14.25 (9.90, 19.46) | 12.58 (9.05, 16.68) | 13.00 (9.78, 16.78) |
| MRT | Mauritania | 92.11 | 116.70 (83.29, 156.10) | 102.90 (75.59, 135.10) | 106.10 (85.82, 130.50) | 14.68 (10.15, 20.11) | 12.94 (9.27, 17.20) | 13.38 (10.03, 17.31) |
| KGZ | Kyrgyz Republic | 96.65 | 122.70 (87.30, 164.70) | 108.10 (79.14, 142.40) | 111.50 (89.98, 137.40) | 15.11 (10.40, 20.76) | 13.30 (9.49, 17.73) | 13.75 (10.28, 17.84) |
| COM | Comoros | 102.2 | 130.20 (92.23, 175.30) | 114.50 (83.51, 151.30) | 118.20 (95.09, 145.90) | 15.62 (10.71, 21.55) | 13.73 (9.76, 18.37) | 14.20 (10.58, 18.48) |
| LBR | Liberia | 103.9 | 132.50 (93.74, 178.60) | 116.50 (84.84, 154.10) | 120.20 (96.65, 148.60) | 15.78 (10.80, 21.79) | 13.86 (9.84, 18.56) | 14.34 (10.67, 18.68) |
| SLB | Solomon Islands | 111.4 | 142.50 (100.30, 192.90) | 125.00 (90.62, 166.00) | 129.10 (103.40, 160.00) | 16.44 (11.19, 22.81) | 14.41 (10.18, 19.38) | 14.92 (11.05, 19.51) |
| KHM | Cambodia | 114.6 | 146.80 (103.10, 199.10) | 128.70 (93.11, 171.20) | 132.90 (106.40, 165.00) | 16.72 (11.35, 23.24) | 14.65 (10.32, 19.73) | 15.17 (11.21, 19.86) |
| LSO | Lesotho | 122.6 | 157.80 (110.20, 214.90) | 138.10 (99.38, 184.40) | 142.60 (113.70, 177.60) | 17.42 (11.75, 24.32) | 15.23 (10.67, 20.60) | 15.77 (11.61, 20.73) |
| IDN | Indonesia | 131.8 | 170.20 (118.20, 232.80) | 148.60 (106.50, 199.30) | 153.60 (122.10, 191.80) | 18.18 (12.19, 25.50) | 15.86 (11.05, 21.55) | 16.44 (12.04, 21.69) |
| LKA | Sri Lanka | 134.2 | 173.60 (120.40, 237.70) | 151.50 (108.40, 203.30) | 156.50 (124.30, 195.70) | 18.38 (12.31, 25.82) | 16.03 (11.16, 21.80) | 16.62 (12.15, 21.94) |
| EGY | Egypt, Arab Rep. | 141.4 | 183.40 (126.70, 251.90) | 159.80 (113.90, 215.20) | 165.20 (130.90, 207.00) | 18.96 (12.64, 26.73) | 16.51 (11.45, 22.53) | 17.12 (12.48, 22.68) |
| VUT | Vanuatu | 143.4 | 186.20 (128.40, 255.90) | 162.20 (115.40, 218.50) | 167.60 (132.70, 210.20) | 19.13 (12.74, 26.98) | 16.65 (11.53, 22.73) | 17.26 (12.57, 22.88) |
| TLS | Timor-Leste | 144.2 | 187.20 (129.10, 257.50) | 163.10 (116.00, 219.80) | 168.60 (133.40, 211.40) | 19.19 (12.77, 27.08) | 16.70 (11.56, 22.81) | 17.32 (12.61, 22.96) |
| BTN | Bhutan | 162.2 | 212.00 (144.80, 293.60) | 184.00 (129.80, 249.70) | 190.30 (149.80, 239.90) | 20.58 (13.56, 29.27) | 17.85 (12.25, 24.56) | 18.53 (13.38, 24.72) |
| STP | Sao Tome and Principe | 171.1 | 224.20 (152.60, 311.50) | 194.30 (136.60, 264.50) | 201.10 (157.80, 254.10) | 21.25 (13.94, 30.32) | 18.40 (12.57, 25.39) | 19.10 (13.75, 25.57) |
| VEN | Venezuela, RB | 186 | 244.90 (165.60, 342.00) | 211.80 (148.00, 289.60) | 219.20 (171.40, 278.00) | 22.34 (14.55, 32.06) | 19.30 (13.10, 26.77) | 20.05 (14.35, 26.95) |
| UZB | Uzbekistan | 192.1 | 253.40 (170.90, 354.70) | 219.00 (152.70, 300.00) | 226.70 (176.90, 287.90) | 22.78 (14.79, 32.75) | 19.66 (13.31, 27.32) | 20.43 (14.59, 27.51) |
| PHL | Philippines | 194.1 | 256.10 (172.60, 358.60) | 221.20 (154.10, 303.30) | 229.00 (178.70, 291.00) | 22.91 (14.87, 32.97) | 19.77 (13.38, 27.49) | 20.54 (14.66, 27.68) |
| VNM | Viet Nam | 197.2 | 324.40 (86.80, 835.10) | 232.60 (73.83, 594.80) | 301.20 (122.70, 680.30) | 33.47 (6.05, 110.00) | 22.95 (5.42, 67.37) | 30.94 (7.54, 91.02) |
| FJI | Fiji | 212.9 | 282.50 (188.90, 397.70) | 243.30 (168.40, 335.30) | 252.00 (195.70, 321.60) | 24.22 (15.59, 35.06) | 20.85 (14.00, 29.14) | 21.67 (15.37, 29.35) |
| NIC | Nicaragua | 216.6 | 287.60 (192.10, 405.40) | 247.60 (171.20, 341.60) | 256.50 (199.10, 327.50) | 24.47 (15.73, 35.46) | 21.05 (14.12, 29.45) | 21.89 (15.51, 29.66) |
| MAR | Morocco | 232.2 | 309.60 (205.70, 438.30) | 266.10 (183.00, 368.50) | 275.80 (213.30, 353.20) | 25.51 (16.30, 37.15) | 21.91 (14.61, 30.78) | 22.79 (16.07, 31.00) |
| DZA | Algeria | 233.4 | 311.20 (206.70, 440.70) | 267.40 (183.90, 370.50) | 277.10 (214.30, 355.00) | 25.59 (16.34, 37.27) | 21.97 (14.65, 30.87) | 22.85 (16.11, 31.10) |
| GNQ | Equatorial Guinea | 233.7 | 311.70 (207.00, 441.30) | 267.80 (184.10, 371.00) | 277.50 (214.60, 355.50) | 25.61 (16.36, 37.30) | 21.98 (14.66, 30.90) | 22.87 (16.12, 31.12) |
| KIR | Kiribati | 238.1 | 317.90 (210.80, 450.70) | 273.00 (187.40, 378.70) | 283.00 (218.60, 362.80) | 25.90 (16.51, 37.77) | 22.22 (14.79, 31.27) | 23.12 (16.28, 31.49) |
| HND | Honduras | 249.2 | 333.50 (220.40, 474.10) | 286.00 (195.70, 397.70) | 296.50 (228.50, 380.90) | 26.61 (16.90, 38.92) | 22.80 (15.13, 32.17) | 23.73 (16.66, 32.41) |
| GAB | Gabon | 253 | 338.90 (223.70, 482.20) | 290.50 (198.60, 404.40) | 301.20 (232.00, 387.20) | 26.85 (17.03, 39.31) | 23.00 (15.24, 32.48) | 23.94 (16.79, 32.72) |
| BOL | Bolivia | 255 | 341.80 (225.50, 486.60) | 293.00 (200.20, 407.90) | 303.80 (233.80, 390.60) | 26.99 (17.10, 39.53) | 23.11 (15.30, 32.65) | 24.05 (16.86, 32.89) |
| LBN | Lebanon | 260.8 | 350.00 (230.40, 498.90) | 299.80 (204.50, 417.90) | 310.90 (239.10, 400.20) | 27.35 (17.30, 40.12) | 23.40 (15.47, 33.11) | 24.36 (17.05, 33.35) |
| IRN | Iran, Islamic Rep. | 267.1 | 358.90 (235.80, 512.30) | 307.20 (209.20, 428.80) | 318.60 (244.70, 410.50) | 27.74 (17.51, 40.75) | 23.72 (15.65, 33.61) | 24.70 (17.26, 33.85) |
| CPV | Cabo Verde | 290.1 | 391.70 (255.70, 561.90) | 334.50 (226.40, 469.10) | 347.00 (265.50, 448.80) | 29.15 (18.27, 43.06) | 24.87 (16.30, 35.40) | 25.91 (18.00, 35.67) |
| WSM | Samoa | 292.8 | 395.50 (258.00, 567.70) | 337.70 (228.40, 473.90) | 350.30 (267.90, 453.30) | 29.31 (18.36, 43.32) | 25.00 (16.38, 35.61) | 26.04 (18.09, 35.88) |
| SWZ | Eswatini | 296 | 400.10 (260.80, 574.80) | 341.50 (230.80, 479.50) | 354.30 (270.80, 458.70) | 29.50 (18.46, 43.64) | 25.15 (16.47, 35.86) | 26.21 (18.19, 36.13) |
| SUR | Suriname | 297 | 401.60 (261.70, 576.90) | 342.70 (231.60, 481.30) | 355.60 (271.70, 460.30) | 29.56 (18.49, 43.73) | 25.20 (16.49, 35.93) | 26.26 (18.22, 36.20) |
| AZE | Azerbaijan | 299.8 | 405.50 (264.10, 582.90) | 346.00 (233.60, 486.20) | 359.00 (274.20, 465.00) | 29.73 (18.58, 44.00) | 25.34 (16.57, 36.14) | 26.40 (18.31, 36.42) |
| BLZ | Belize | 309.4 | 419.20 (272.30, 603.80) | 357.40 (240.80, 503.10) | 370.90 (282.80, 481.00) | 30.30 (18.88, 44.93) | 25.80 (16.83, 36.87) | 26.88 (18.60, 37.15) |
| TUN | Tunisia | 317.7 | 431.10 (279.50, 621.90) | 367.20 (247.00, 517.70) | 381.20 (290.30, 494.90) | 30.78 (19.14, 45.73) | 26.19 (17.05, 37.49) | 27.30 (18.85, 37.77) |
| THA | Thailand | 326.5 | 679.30 (330.40, 1392) | 480.90 (321.00, 715.80) | 592.40 (287.70, 1193) | 53.44 (20.27, 126.40) | 36.91 (21.52, 61.02) | 46.78 (17.54, 107.40) |
| IRQ | Iraq | 333.3 | 453.50 (292.90, 656.10) | 385.70 (258.50, 545.30) | 400.50 (304.30, 521.10) | 31.68 (19.62, 47.21) | 26.91 (17.46, 38.63) | 28.06 (19.32, 38.93) |
| JOR | Jordan | 342.1 | 466.20 (300.40, 675.50) | 396.20 (265.10, 561.00) | 411.40 (312.20, 536.00) | 32.17 (19.88, 48.03) | 27.32 (17.68, 39.27) | 28.49 (19.58, 39.57) |
| PSE | West Bank and Gaza | 345.7 | 471.30 (303.50, 683.40) | 400.50 (267.70, 567.30) | 415.90 (315.40, 542.00) | 32.38 (19.99, 48.36) | 27.48 (17.77, 39.53) | 28.66 (19.68, 39.83) |
| TON | Tonga | 357.5 | 488.40 (313.70, 709.60) | 414.60 (276.50, 588.50) | 430.60 (326.10, 562.10) | 33.04 (20.34, 49.46) | 28.01 (18.07, 40.38) | 29.22 (20.02, 40.69) |
| MNG | Mongolia | 359.6 | 491.40 (315.50, 714.20) | 417.10 (278.00, 592.20) | 433.20 (328.00, 565.60) | 33.15 (20.40, 49.65) | 28.11 (18.12, 40.52) | 29.32 (20.08, 40.84) |
| GTM | Guatemala | 392 | 538.30 (343.20, 786.40) | 455.80 (301.90, 650.30) | 473.60 (357.00, 620.60) | 34.91 (21.32, 52.57) | 29.52 (18.91, 42.78) | 30.82 (20.99, 43.12) |
| NAM | Namibia | 397.1 | 545.70 (347.60, 797.80) | 461.90 (305.70, 659.40) | 480.00 (361.60, 629.30) | 35.18 (21.46, 53.02) | 29.74 (19.03, 43.13) | 31.05 (21.12, 43.47) |
| UKR | Ukraine | 410.5 | 541.00 (138.00, 1434) | 497.90 (88.47, 1497) | 431.30 (167.70, 931.10) | 38.40 (9.61, 107.30) | 33.97 (6.91, 99.00) | 30.35 (11.98, 70.02) |
| PER | Peru | 445.5 | 616.30 (389.00, 907.20) | 519.90 (341.20, 747.10) | 540.60 (404.90, 712.30) | 37.69 (22.76, 57.23) | 31.76 (20.14, 46.37) | 33.18 (22.40, 46.74) |
| MYS | Malaysia | 450.3 | 623.30 (393.10, 918.10) | 525.70 (344.80, 755.90) | 546.60 (409.30, 720.60) | 37.94 (22.89, 57.64) | 31.95 (20.24, 46.68) | 33.38 (22.52, 47.06) |
| MDA | Moldova | 451.7 | 625.40 (394.30, 921.30) | 527.30 (345.80, 758.40) | 548.30 (410.50, 723.00) | 38.01 (22.93, 57.76) | 32.01 (20.27, 46.77) | 33.44 (22.56, 47.15) |
| VCT | St. Vincent and the Grenadines | 469.2 | 651.00 (409.20, 961.20) | 548.30 (358.60, 790.30) | 570.30 (426.10, 753.20) | 38.88 (23.38, 59.23) | 32.71 (20.66, 47.91) | 34.18 (23.00, 48.29) |
| LBY | Libya | 470.4 | 652.70 (410.20, 963.90) | 549.80 (359.40, 792.50) | 571.80 (427.20, 755.20) | 38.94 (23.41, 59.33) | 32.76 (20.68, 47.98) | 34.23 (23.03, 48.37) |
| DOM | Dominican Republic | 488.9 | 679.80 (425.90, 1006) | 572.00 (372.90, 826.30) | 595.00 (443.70, 787.20) | 39.85 (23.87, 60.87) | 33.49 (21.08, 49.16) | 35.00 (23.49, 49.56) |
| BWA | Botswana | 489.6 | 680.90 (426.60, 1008) | 572.90 (373.40, 827.60) | 595.90 (444.30, 788.50) | 39.88 (23.89, 60.93) | 33.51 (21.09, 49.20) | 35.03 (23.50, 49.61) |
| KAZ | Kazakhstan | 489.7 | 682.00 (176.30, 1822) | 646.00 (115.90, 2119) | 545.90 (227.70, 1202) | 40.58 (9.54, 112.80) | 36.40 (7.04, 115.80) | 32.16 (11.99, 75.69) |
| DMA | Dominica | 492.5 | 685.10 (429.00, 1015) | 576.30 (375.50, 832.80) | 599.50 (446.90, 793.40) | 40.02 (23.96, 61.17) | 33.63 (21.15, 49.38) | 35.15 (23.57, 49.79) |
| SLV | El Salvador | 500.7 | 633.20 (121.90, 1804) | 446.20 (127.30, 1206) | 563.60 (128.40, 1634) | 41.70 (5.47, 155.30) | 27.78 (5.74, 87.86) | 37.70 (5.20, 138.30) |
| GUY | Guyana | 505 | 703.60 (439.70, 1043) | 591.50 (384.70, 855.90) | 615.30 (458.10, 815.30) | 40.63 (24.27, 62.20) | 34.11 (21.42, 50.17) | 35.67 (23.88, 50.59) |
| FSM | Micronesia, Fed. Sts. | 507.8 | 707.70 (442.00, 1050) | 594.80 (386.70, 861.00) | 618.80 (460.60, 820.10) | 40.77 (24.34, 62.42) | 34.22 (21.48, 50.35) | 35.78 (23.95, 50.76) |
| ECU | Ecuador | 509.5 | 710.20 (443.50, 1054) | 596.80 (387.90, 864.10) | 621.00 (462.10, 823.00) | 40.85 (24.38, 62.56) | 34.29 (21.51, 50.45) | 35.85 (23.99, 50.87) |
| JAM | Jamaica | 520.2 | 725.90 (452.60, 1078) | 609.70 (395.70, 883.80) | 634.40 (471.60, 841.60) | 41.36 (24.64, 63.43) | 34.69 (21.73, 51.11) | 36.28 (24.24, 51.54) |
| GRD | Grenada | 526.4 | 735.10 (457.90, 1093) | 617.20 (400.20, 895.30) | 642.30 (477.20, 852.50) | 41.66 (24.79, 63.93) | 34.93 (21.86, 51.50) | 36.53 (24.39, 51.93) |
| PRY | Paraguay | 526.5 | 735.30 (458.00, 1093) | 617.30 (400.30, 895.50) | 642.40 (477.30, 852.70) | 41.66 (24.80, 63.94) | 34.93 (21.86, 51.51) | 36.53 (24.39, 51.93) |
| ZAF | South Africa | 536.6 | 1169 (391.80, 2836) | 960.40 (425.10, 2081) | 1007 (495.10, 1955) | 66.01 (19.15, 170.10) | 52.69 (22.95, 110.70) | 56.90 (23.60, 123.20) |
| TUR | Turkiye | 547.8 | 766.80 (476.10, 1143) | 643.00 (415.80, 934.90) | 669.30 (496.30, 889.90) | 42.66 (25.31, 65.65) | 35.73 (22.29, 52.81) | 37.38 (24.89, 53.25) |
| GEO | Georgia | 554.3 | 776.40 (481.60, 1158) | 650.90 (420.50, 946.90) | 677.50 (502.10, 901.30) | 42.97 (25.46, 66.16) | 35.98 (22.42, 53.20) | 37.64 (25.04, 53.65) |
| BLR | Belarus | 558.2 | 782.20 (484.90, 1167) | 655.60 (423.40, 954.20) | 682.40 (505.60, 908.10) | 43.15 (25.55, 66.47) | 36.12 (22.50, 53.44) | 37.79 (25.13, 53.88) |
| COL | Colombia | 571.2 | 4994 (933.40, 13810) | 3021 (1545, 5632) | 4833 (2625, 8908) | 336.60 (56.22, 1011) | 197.60 (99.83, 385.10) | 330.10 (134.70, 742.30) |
| TKM | Turkmenistan | 581.4 | 816.50 (504.60, 1221) | 683.60 (440.10, 997.20) | 711.70 (526.20, 948.80) | 44.21 (26.09, 68.29) | 36.97 (22.96, 54.82) | 38.69 (25.66, 55.28) |
| ALB | Albania | 590.6 | 830.30 (512.40, 1243) | 694.80 (446.80, 1014) | 723.40 (534.50, 965.00) | 44.63 (26.30, 69.01) | 37.30 (23.13, 55.37) | 39.04 (25.86, 55.83) |
| MUS | Mauritius | 616.2 | 868.30 (534.10, 1303) | 725.80 (465.30, 1062) | 755.80 (557.20, 1010) | 45.78 (26.88, 70.98) | 38.22 (23.62, 56.86) | 40.01 (26.43, 57.35) |
| MKD | North Macedonia | 639.4 | 903.00 (553.80, 1358) | 753.90 (482.10, 1106) | 785.30 (577.90, 1051) | 46.81 (27.39, 72.74) | 39.03 (24.06, 58.20) | 40.87 (26.93, 58.70) |
| LCA | St. Lucia | 641.1 | 905.40 (555.20, 1362) | 755.90 (483.30, 1109) | 787.40 (579.40, 1054) | 46.88 (27.43, 72.86) | 39.09 (24.09, 58.30) | 40.93 (26.97, 58.79) |
| OMN | Oman | 732 | 1042 (632.10, 1579) | 866.50 (548.60, 1280) | 903.10 (660.20, 1216) | 50.77 (29.36, 79.57) | 42.17 (25.71, 63.37) | 44.19 (28.85, 63.92) |
| BRN | Brunei Darussalam | 733 | 1043 (632.90, 1582) | 867.70 (549.30, 1282) | 904.40 (661.10, 1218) | 50.81 (29.38, 79.64) | 42.20 (25.73, 63.42) | 44.23 (28.87, 63.98) |
| MEX | Mexico | 761 | 4484 (879.60, 13610) | 5905 (2429, 13720) | 3040 (1285, 6639) | 241.30 (20.48, 924.60) | 304.90 (57.98, 922.60) | 163.90 (24.52, 500.40) |
| CHN | China | 763.4 | 1839 (768.50, 3647) | 1487 (849.20, 2392) | 1582 (932.80, 2591) | 122.80 (50.40, 246.20) | 97.70 (67.16, 144.10) | 107.20 (57.85, 195.30) |
| SYC | Seychelles | 766.3 | 1093 (661.00, 1662) | 908.30 (573.20, 1345) | 946.80 (690.60, 1277) | 52.18 (30.06, 82.02) | 43.28 (26.30, 65.22) | 45.37 (29.53, 65.79) |
| ARM | Armenia | 767 | 1094 (661.60, 1664) | 909.10 (573.60, 1347) | 947.70 (691.20, 1279) | 52.21 (30.07, 82.07) | 43.30 (26.31, 65.26) | 45.40 (29.55, 65.83) |
| BIH | Bosnia and Herzegovina | 772.9 | 1103 (666.60, 1678) | 916.30 (577.90, 1358) | 955.30 (696.50, 1289) | 52.45 (30.19, 82.49) | 43.49 (26.41, 65.57) | 45.60 (29.66, 66.15) |
| ATG | Antigua and Barbuda | 841.2 | 1207 (724.20, 1845) | 999.80 (626.70, 1489) | 1043 (757.10, 1412) | 55.19 (31.53, 87.26) | 45.65 (27.53, 69.16) | 47.89 (30.97, 69.78) |
| MHL | Marshall Islands | 894.2 | 1287 (768.70, 1975) | 1065 (664.30, 1591) | 1111 (804.00, 1508) | 57.25 (32.53, 90.87) | 47.27 (28.38, 71.87) | 49.61 (31.95, 72.52) |
| SRB | Serbia | 983.7 | 1424 (843.80, 2197) | 1174 (727.80, 1764) | 1226 (883.10, 1672) | 60.62 (34.16, 96.80) | 49.92 (29.74, 76.32) | 52.42 (33.54, 77.01) |
| RUS | Russian Federation | 1003 | 1454 (860.30, 2246) | 1199 (741.70, 1802) | 1251 (900.40, 1708) | 61.35 (34.51, 98.08) | 50.49 (30.03, 77.27) | 53.02 (33.88, 77.98) |
| BRA | Brazil | 1010 | 736.60 (126.50, 2278) | 931.10 (98.08, 3548) | 756.20 (129.20, 2426) | 24.50 (4.00, 86.67) | 29.16 (3.12, 111.50) | 26.20 (3.54, 97.00) |
| ROU | Romania | 1051 | 1738 (459.70, 4531) | 1571 (306.70, 5011) | 1370 (586.10, 2934) | 70.17 (15.57, 203.10) | 60.54 (11.31, 190.70) | 55.40 (19.39, 134.10) |
| MNE | Montenegro | 1148 | 1676 (981.20, 2610) | 1377 (843.40, 2085) | 1438 (1028, 1974) | 66.51 (36.97, 107.20) | 54.52 (32.08, 84.09) | 57.30 (36.29, 84.87) |
| MDV | Maldives | 1159 | 1694 (990.60, 2638) | 1390 (851.40, 2108) | 1452 (1038, 1995) | 66.90 (37.15, 107.90) | 54.83 (32.24, 84.61) | 57.63 (36.47, 85.40) |
| CRI | Costa Rica | 1163 | 1701 (994.20, 2649) | 1396 (854.40, 2116) | 1458 (1042, 2002) | 67.05 (37.22, 108.20) | 54.94 (32.30, 84.81) | 57.75 (36.54, 85.60) |
| BHR | Bahrain | 1176 | 1720 (1005, 2681) | 1411 (863.20, 2141) | 1474 (1053, 2026) | 67.48 (37.43, 109.00) | 55.28 (32.47, 85.38) | 58.11 (36.74, 86.18) |
| CUB | Cuba | 1199 | 1756 (1024, 2741) | 1440 (879.70, 2187) | 1505 (1073, 2069) | 68.29 (37.81, 110.40) | 55.91 (32.79, 86.45) | 58.78 (37.11, 87.26) |
| BGR | Bulgaria | 1258 | 1847 (1073, 2891) | 1513 (920.70, 2303) | 1581 (1125, 2178) | 70.27 (38.74, 114.00) | 57.45 (33.57, 89.08) | 60.42 (38.02, 89.92) |
| TTO | Trinidad and Tobago | 1276 | 1876 (1089, 2938) | 1536 (933.60, 2340) | 1605 (1141, 2213) | 70.89 (39.04, 115.10) | 57.93 (33.81, 89.90) | 60.93 (38.31, 90.75) |
| BRB | Barbados | 1286 | 1892 (1097, 2964) | 1548 (940.70, 2360) | 1618 (1150, 2232) | 71.23 (39.19, 115.70) | 58.20 (33.94, 90.35) | 61.21 (38.46, 91.20) |
| KNA | St. Kitts and Nevis | 1295 | 1905 (1104, 2986) | 1559 (946.60, 2377) | 1629 (1157, 2247) | 71.51 (39.32, 116.20) | 58.41 (34.05, 90.72) | 61.44 (38.59, 91.58) |
| HUN | Hungary | 1420 | 2100 (1208, 3309) | 1714 (1034, 2627) | 1791 (1267, 2482) | 75.58 (41.23, 123.50) | 61.58 (35.63, 96.13) | 64.80 (40.44, 97.06) |
| ARG | Argentina | 1457 | 2947 (661.50, 8294) | 2376 (994.70, 5450) | 2489 (654.50, 6578) | 107.60 (18.26, 341.10) | 82.43 (31.25, 193.60) | 91.83 (17.09, 279.40) |
| HRV | Croatia | 1548 | 2301 (1315, 3646) | 1874 (1123, 2886) | 1959 (1380, 2725) | 79.62 (43.10, 130.80) | 64.70 (37.18, 101.50) | 68.13 (42.27, 102.50) |
| PAN | Panama | 1558 | 2146 (404.60, 6439) | 1424 (407.80, 3940) | 1851 (400.10, 5613) | 83.92 (9.83, 313.50) | 52.97 (9.55, 165.20) | 73.72 (8.84, 277.40) |
| POL | Poland | 1578 | 1973 (527.80, 4959) | 1984 (373.60, 6853) | 1467 (758.50, 2752) | 56.49 (13.25, 151.00) | 53.66 (10.01, 175.70) | 42.25 (17.47, 87.76) |
| LVA | Latvia | 1645 | 2454 (1395, 3902) | 1995 (1190, 3082) | 2086 (1465, 2909) | 82.58 (44.46, 136.20) | 66.99 (38.31, 105.50) | 70.57 (43.60, 106.50) |
| KWT | Kuwait | 1679 | 2508 (1423, 3991) | 2037 (1214, 3151) | 2131 (1495, 2973) | 83.60 (44.93, 138.00) | 67.77 (38.69, 106.80) | 71.40 (44.05, 107.90) |
| PLW | Palau | 1684 | 2515 (1427, 4004) | 2043 (1217, 3160) | 2137 (1499, 2982) | 83.74 (44.99, 138.30) | 67.88 (38.75, 107.00) | 71.52 (44.12, 108.10) |
| TUV | Tuvalu | 1724 | 2578 (1460, 4109) | 2092 (1244, 3241) | 2189 (1534, 3058) | 84.92 (45.53, 140.40) | 68.79 (39.19, 108.60) | 72.49 (44.64, 109.70) |
| CHL | Chile | 1737 | 2599 (1471, 4144) | 2108 (1253, 3268) | 2206 (1545, 3083) | 85.30 (45.71, 141.10) | 69.09 (39.34, 109.10) | 72.80 (44.82, 110.20) |
| QAT | Qatar | 1804 | 2706 (1527, 4325) | 2193 (1300, 3407) | 2295 (1604, 3213) | 87.29 (46.61, 144.80) | 70.62 (40.09, 111.80) | 74.43 (45.70, 112.90) |
| SVK | Slovak Republic | 1813 | 2721 (1535, 4349) | 2205 (1306, 3425) | 2307 (1612, 3230) | 87.56 (46.73, 145.30) | 70.83 (40.19, 112.10) | 74.65 (45.82, 113.20) |
| SAU | Saudi Arabia | 1825 | 2739 (1544, 4381) | 2220 (1314, 3449) | 2323 (1623, 3253) | 87.90 (46.89, 145.90) | 71.09 (40.32, 112.60) | 74.93 (45.97, 113.70) |
| GRC | Greece | 1963 | 2958 (1658, 4750) | 2392 (1409, 3732) | 2504 (1743, 3517) | 91.82 (48.66, 153.10) | 74.10 (41.78, 117.80) | 78.14 (47.70, 119.00) |
| LTU | Lithuania | 2044 | 3088 (1725, 4971) | 2494 (1465, 3900) | 2611 (1814, 3675) | 94.09 (49.69, 157.30) | 75.85 (42.63, 120.90) | 80.01 (48.70, 122.10) |
| URY | Uruguay | 2076 | 3139 (1752, 5058) | 2534 (1487, 3966) | 2654 (1842, 3737) | 94.98 (50.09, 158.90) | 76.53 (42.96, 122.10) | 80.73 (49.09, 123.30) |
| EST | Estonia | 2258 | 3432 (1902, 5557) | 2764 (1611, 4346) | 2895 (2001, 4091) | 99.91 (52.29, 168.00) | 80.30 (44.77, 128.70) | 84.76 (51.24, 130.00) |
| BHS | Bahamas, The | 2262 | 3438 (1905, 5567) | 2769 (1614, 4354) | 2900 (2005, 4099) | 100.00 (52.34, 168.20) | 80.38 (44.81, 128.90) | 84.84 (51.28, 130.20) |
| NRU | Nauru | 2292 | 3487 (1930, 5650) | 2807 (1634, 4417) | 2940 (2031, 4157) | 100.80 (52.69, 169.70) | 80.99 (45.10, 129.90) | 85.49 (51.63, 131.30) |
| ARE | United Arab Emirates | 2402 | 3663 (2020, 5951) | 2944 (1709, 4645) | 3085 (2126, 4371) | 103.70 (53.97, 175.00) | 83.18 (46.15, 133.80) | 87.82 (52.87, 135.20) |
| TWN | Taiwan | 2522 | 5080 (1103, 15070) | 4386 (743.40, 14660) | 3969 (1374, 10270) | 155.60 (23.02, 524.90) | 125.80 (18.37, 464.40) | 123.40 (25.76, 381.30) |
| CZE | Czechia | 2671 | 4100 (2241, 6702) | 3286 (1892, 5213) | 3444 (2361, 4902) | 110.50 (56.99, 187.80) | 88.40 (48.63, 143.10) | 93.40 (55.82, 144.60) |
| PRT | Portugal | 2762 | 4248 (2316, 6958) | 3401 (1953, 5407) | 3565 (2440, 5082) | 112.80 (57.98, 192.10) | 90.12 (49.44, 146.10) | 95.23 (56.78, 147.60) |
| CYP | Cyprus | 2899 | 4472 (2428, 7344) | 3575 (2046, 5698) | 3749 (2559, 5354) | 116.10 (59.43, 198.30) | 92.65 (50.63, 150.60) | 97.94 (58.20, 152.20) |
| SVN | Slovenia | 3038 | 4698 (2541, 7737) | 3751 (2139, 5994) | 3934 (2679, 5630) | 119.40 (60.87, 204.60) | 95.16 (51.81, 155.10) | 100.60 (59.60, 156.80) |
| KOR | Korea, Rep. | 3044 | 4708 (2546, 7754) | 3759 (2143, 6007) | 3942 (2685, 5642) | 119.60 (60.93, 204.80) | 95.27 (51.86, 155.30) | 100.70 (59.66, 157.00) |
| ESP | Spain | 3107 | 6799 (2042, 16370) | 5307 (2791, 10110) | 5666 (2879, 10880) | 169.10 (39.57, 445.20) | 126.30 (59.98, 240.40) | 141.90 (46.84, 329.30) |
| ITA | Italy | 3283 | 6993 (1738, 18450) | 4992 (2873, 8478) | 5828 (1697, 14690) | 202.00 (41.65, 579.10) | 139.10 (74.08, 256.40) | 170.40 (40.32, 503.20) |
| AND | Andorra | 3605 | 5633 (3005, 9367) | 4475 (2520, 7218) | 4696 (3171, 6770) | 132.40 (66.45, 229.20) | 105.00 (56.36, 172.80) | 111.10 (65.03, 174.60) |
| MLT | Malta | 3623 | 5663 (3019, 9420) | 4499 (2532, 7257) | 4721 (3187, 6807) | 132.80 (66.63, 230.00) | 105.30 (56.51, 173.30) | 111.40 (65.20, 175.20) |
| JPN | Japan | 3638 | 8612 (3123, 19630) | 7375 (1496, 21980) | 7767 (4030, 14270) | 241.20 (90.58, 527.40) | 200.40 (46.30, 556.20) | 219.10 (116.60, 407.40) |
| SGP | Singapore | 3922 | 6158 (3262, 10290) | 4881 (2731, 7907) | 5124 (3445, 7412) | 139.30 (69.38, 242.30) | 110.20 (58.75, 182.10) | 116.70 (67.88, 184.10) |
| SMR | San Marino | 3924 | 6161 (3264, 10300) | 4883 (2732, 7912) | 5126 (3447, 7416) | 139.30 (69.40, 242.40) | 110.20 (58.76, 182.20) | 116.70 (67.90, 184.20) |
| ISR | Israel | 3928 | 6168 (3267, 10310) | 4888 (2735, 7921) | 5132 (3450, 7425) | 139.40 (69.44, 242.60) | 110.30 (58.79, 182.30) | 116.80 (67.94, 184.30) |
| HKG | Hong Kong | 4257 | 5594 (993.40, 17440) | 4912 (696.40, 17660) | 4064 (1096, 11240) | 128.90 (14.96, 480.00) | 108.00 (11.87, 414.50) | 94.93 (14.69, 314.70) |
| NZL | New Zealand | 4997 | 6290 (1524, 16970) | 6441 (1113, 22300) | 4427 (2236, 8603) | 98.85 (19.30, 286.50) | 94.27 (14.51, 301.50) | 69.82 (24.61, 153.00) |
| FRA | France | 5149 | 8218 (4257, 13950) | 6462 (3544, 10620) | 6791 (4504, 9938) | 164.10 (79.77, 290.30) | 128.80 (67.18, 216.20) | 136.60 (77.99, 218.60) |
| GBR | United Kingdom | 5407 | 8511 (2649, 20300) | 7709 (3990, 14850) | 6085 (3591, 10150) | 168.20 (43.70, 420.90) | 146.50 (77.82, 268.80) | 121.10 (50.94, 237.10) |
| FIN | Finland | 5515 | 7155 (1423, 21590) | 4206 (1618, 10060) | 6021 (1351, 17930) | 161.20 (20.35, 634.70) | 90.24 (21.69, 267.60) | 139.60 (17.93, 555.30) |
| BEL | Belgium | 5931 | 11840 (5306, 25370) | 10520 (1935, 33820) | 10300 (2935, 27160) | 245.80 (59.85, 675.60) | 206.80 (31.34, 688.90) | 217.40 (36.62, 686.50) |
| CAN | Canada | 6187 | 12220 (5447, 27050) | 9931 (2028, 30800) | 8171 (3533, 16990) | 243.60 (61.89, 671.80) | 186.00 (31.92, 597.80) | 164.20 (38.58, 442.10) |
| SWE | Sweden | 6206 | 10020 (5110, 17180) | 7835 (4236, 13010) | 8239 (5413, 12150) | 183.70 (87.78, 328.60) | 143.30 (73.64, 243.10) | 152.20 (85.78, 245.90) |
| AUT | Austria | 6268 | 8968 (1999, 24960) | 8124 (1429, 29320) | 6399 (2686, 14700) | 149.30 (27.67, 457.70) | 127.90 (21.93, 443.80) | 106.90 (33.95, 272.60) |
| NLD | Netherlands | 6372 | 9521 (3181, 22860) | 9163 (1669, 28690) | 7993 (3153, 18630) | 167.80 (55.23, 425.10) | 153.40 (29.89, 478.80) | 146.20 (44.22, 372.80) |
| DEU | Germany | 6395 | 4246 (1446, 8530) | 4027 (2130, 7254) | 4486 (2696, 7458) | 68.43 (29.27, 135.80) | 65.52 (37.33, 110.90) | 76.09 (36.71, 147.10) |
| DNK | Denmark | 6511 | 10540 (5355, 18120) | 8231 (4435, 13700) | 8657 (5674, 12790) | 189.10 (89.96, 339.20) | 147.30 (75.40, 250.60) | 156.50 (87.90, 253.50) |
| IRL | Ireland | 6878 | 11170 (5650, 19270) | 8710 (4674, 14540) | 9163 (5989, 13570) | 195.40 (92.53, 351.80) | 152.00 (77.47, 259.40) | 161.50 (90.39, 262.40) |
| AUS | Australia | 6980 | 10370 (2353, 29610) | 9526 (1640, 32180) | 7603 (3123, 17080) | 166.00 (33.19, 517.90) | 143.60 (23.24, 460.60) | 121.80 (38.23, 312.30) |
| ISL | Iceland | 7065 | 11490 (5800, 19850) | 8954 (4795, 14970) | 9421 (6149, 13970) | 198.60 (93.80, 358.10) | 154.40 (78.49, 263.80) | 164.10 (91.64, 266.90) |
| LUX | Luxembourg | 7511 | 12260 (6158, 21260) | 9538 (5084, 16000) | 10040 (6532, 14920) | 206.10 (96.80, 373.00) | 159.90 (80.90, 274.10) | 170.00 (94.54, 277.40) |
| MCO | Monaco | 8004 | 13120 (6553, 22820) | 10190 (5403, 17150) | 10720 (6953, 15980) | 214.20 (100.00, 389.10) | 165.80 (83.47, 285.30) | 176.40 (97.66, 288.70) |
| NOR | Norway | 8296 | 13630 (6787, 23760) | 10570 (5592, 17830) | 11130 (7203, 16610) | 218.80 (101.90, 398.40) | 169.30 (84.96, 291.80) | 180.10 (99.46, 295.30) |
| LIE | Liechtenstein | 11490 | 19260 (9336, 34190) | 14800 (7638, 25390) | 15590 (9930, 23600) | 266.50 (120.40, 494.70) | 204.20 (99.75, 358.30) | 217.60 (117.40, 362.70) |
| CHE | Switzerland | 11780 | 19770 (9565, 35150) | 15180 (7821, 26080) | 16000 (10180, 24240) | 270.50 (121.90, 502.90) | 207.10 (101.00, 363.90) | 220.70 (118.90, 368.40) |
| USA | United States | 13470 | 29300 (19100, 43380) | 13120 (5080, 28130) | 20630 (13780, 30630) | 519.70 (394.30, 685.20) | 236.00 (88.93, 503.10) | 376.70 (213.60, 622.30) |

**Table S4.** Predicted mean cost of influenza treatment (with 95% posterior intervals), in 195 countries and territories, as predicted by the chosen model, for each age group and treatment setting. All costs presented in 2023 USD.
